# Wearable technology for health monitoring during pregnancy: an observational cross-sectional survey study

**DOI:** 10.1101/2022.01.26.22269158

**Authors:** Colin Wakefield, Lena Yao, Steve Self, Martin G. Frasch

**Author notes:** **Address of correspondence:** Martin G. Frasch, Department of Obstetrics and Gynecology, University of Washington, 1959 NE Pacific St, Box 356460, Seattle, WA 98195.

## Abstract

**Objective:** Telemedicine has advanced to the forefront of healthcare delivery, including maternal-fetal medicine. Smart wearable electrocardiogram (ECG) devices can enable pregnant women to monitor their health and that of their fetuses. Such technology would be a logical extension of the telemedicine ecosystem. However, it is not known how pregnant women perceive the ability to use such technologies.

**Design:** Observational cross-sectional study.

**Setting:** Online survey in the United States in 2019.

**Population:** A representative sample of 507 women aged 18-45 were polled from 45 states.

**Methods:** Study participants were recruited using the SurveyMonkey Audience Polling system and responded virtually.

**Main Outcome Measures:** Women were asked to identify willingness to use a wearable ECG device the size of a patch-sized large band-aid on their abdomen. Ten binary or multiple-choice questions were used to gauge population interest and related demographics towards the usage of a wearable ECG device.

**Results:** 91% of women expecting to become pregnant in the next five years accept wearable ECG technology as a mechanism for increased frequency of monitoring of maternal and fetal health throughout the pregnancy outside the hospital. 78% of women demonstrated a willingness to wear devices day and night or at least during sleep and 42% of women would spend up to $200 on such a device.

**Conclusions:** Even though conducted prior to the COVID-19 pandemic, this study clearly indicates a high degree of readiness of prospective pregnant women for telemedicine with continuous health monitoring of the mother-fetus dyad during the entire antepartum period.

**Funding:** n/a

## Introduction

The COVID-19 pandemic has drastically altered the landscape in which healthcare is delivered. Prior to 2019, telemedicine was pitched as the next frontier. However, little was understood on how it would become integrated into modern healthcare delivery. Over the past two years, patients have become increasingly accepting of receiving medical care remotely.^1^ This represents an exciting social shift that offers enormous opportunities for the improvement of Maternal and Fetal Medicine.

The use of fetal electrocardiogram (ECG) derived from maternal abdominal ECG as a biomarker of fetal well-being offers a prime entry point for the implementation of remote monitoring and telemedicine for this underserved patient population. ECG patterns are currently being investigated as early biomarkers of poor fetal and postnatal development.^2,3^ Studies indicate that in-home stimuli like maternal stress can negatively impact lifelong neurodevelopmental trajectories.^4^ Current practice is only beginning to use ECG as a source of biomarkers intrapartum and not at all antepartum. This leaves minimal time for intervention and correction of fetal development. Applying this technology to the in-home setting will expand the intervention window for providers, improving pregnancy outcomes. Mothers would continuously wear ECG devices monitoring both maternal and fetal ECG and its derivatives, such as heart rate (HR) and HR variability metrics, providing instant on-site and remote access to the health of the mother-fetus dyad. The first step in the implementation of such practices is gauging how perceptive prospective and existing mothers would be to the integration of wearable ECG devices in their daily lives.

## Methods

To gauge their interest in the use of a wearable maternal-fetal ECG device, we used the SurveyMonkey Audience system, from July 31st to August 1st of 2019, to survey 507 female participants across the United States of the ages 18-45, with annual income brackets of $0-$200,000.

SurveyMonkey is an online survey tool used to collect data from individuals across the US population. A pool of over two million people is maintained through an agreement wherein participants agree to take part in a survey, in exchange for SurveyMonkey making a donation of $0.50 to the charity of that individual’s choice. The SurveyMonkey algorithm then randomly assigns participants to surveys in a manner that creates a sample representative of the demographics specified by the investigator. Participants then receive a URL via email from which the survey can be completed on a laptop or mobile device.

### Study ethics approach

Each survey participant was first given the option to opt out of the survey (Question 1). Only if agreed to continue, were the subsequent questions shown. Due to the completely anonymous nature of the data collection and the provided right to opt out of the survey or agree to continue, no IRB approval was obtained prior to the survey’s completion. However, after the completion of the survey and prior to publication of the findings, we consulted the UW IRB. It was determined that the approach taken meets the definition of “minor non-compliance” with IRB approved procedures/UW policies and procedures, because it posed no significant increase in risk or any decrease in benefits to subjects. The IRB ruled that this means that the data collected as part of this research cannot be described as part of a study reviewed by human subject division (IRB). On January 24, 2022, the IRB determined that the corrective actions described in the report are sufficient. No additional actions are required at this time.

### Survey structure

This study was conducted under the parameters of a nationally representative sample containing 500 females ranging 18-45 years of age with an annual income range of $0-200,000. Participants were asked a total of 10 questions. We screened the participants by asking upfront: “Are you planning to be pregnant in the next 5 years?” If no, the questionnaire was stopped, if yes, participants answered four questions to gauge interest in a wearable ECG device as seen in **Figure 1**. When asked question four, participants were given a description of a wearable device the size of a “patch-sized large band-aid” worn on the abdomen for at least 8 continuous hours throughout the day to help doctors ensure they are doing ok during pregnancy when on the go. Demographics were then gathered via five additional questions to identify income, age, location, gender, polling device type distribution among the cohort.

**Figure 1.**
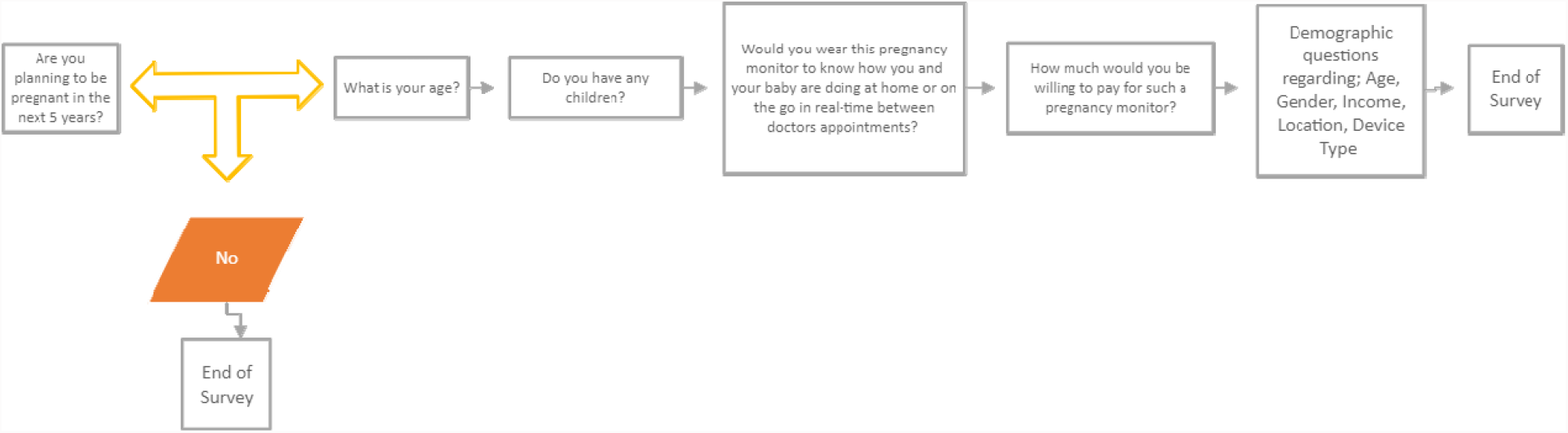
Survey question algorithm.

To achieve adequate power to accurately represent the roughly 60 million US females between the ages of 18-45, we determined 500 participants were needed. In order to gather enough responses for each of the ten questions, SurveyMonkey recruited a total of 527 participants. The response rate among recruited individuals was 96%, with twenty recruited individuals failing to complete all ten questions. The initial screening question had a margin of error of 4.44% due to 507 individuals responding “yes” on “planning to become pregnant in the next 5 years”. Questions 2-5 had a margin of error of 6% due to participants choosing not to answer. Questions 6-10 which were used to gather cohort demographics had a margin of 4.4%. In total, the data collected using the SurveyMonkey Audience System had a confidence level of 95%.

## Results

The cohort represented individuals from forty-five states across the U.S.A., with response density reflecting population densities (Fig. 2). A 97% response rate was achieved because participants were recruited and agreed to complete a survey in principle prior to participation. Individuals had no knowledge of the survey content prior to agreeing to participate. SurveyMonkey randomly assigned participants to the survey until all questions had a response rate of 250. This led to a total recruitment of 527 individuals, twenty of which did not complete the survey and whose response could not be included. Participants make up females 18-45 with incomes ranging $0-200,000 from 45 of the 50 states in the U.S.

**Figure 2.**
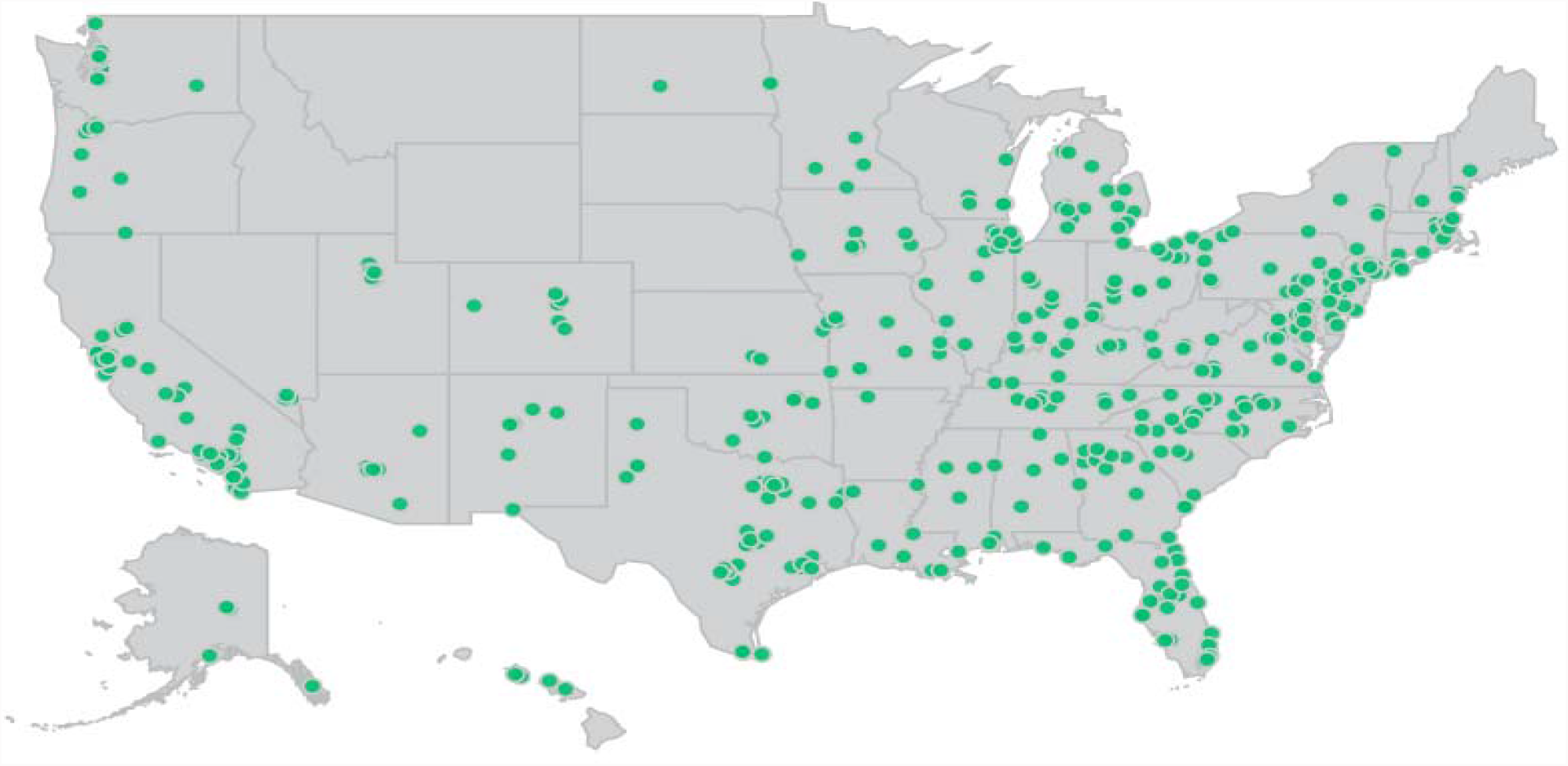
Geographic representation of surveyed participants. The cohort represented individuals from forty-five states across the U.S.A., with response density reflecting population densities

As Table 1 summarizes, a large proportion (43.8%) of participants had already gone through a prior pregnancy. The screening question identified 258 participants, or roughly half of the participants (50.89%) as planning to become pregnant within five years of participating in the study. These participants expressed a willingness to wear an ECG device continuously (46.9%), only while sleeping (31.4%), or only while awake (12.8%) (Fig. 3). Individuals were also polled on their inclinations to spend money on said device, 44.7% were willing to spend up to $100 on such a product and another 41.7%, $100 to $200 (Fig. 3). A subset (9.79%) expressed a willingness to spend up to $300, and an additional 3.83% demonstrated a readiness to spend more than $300 on a wearable ECG device.

**Table 1.**
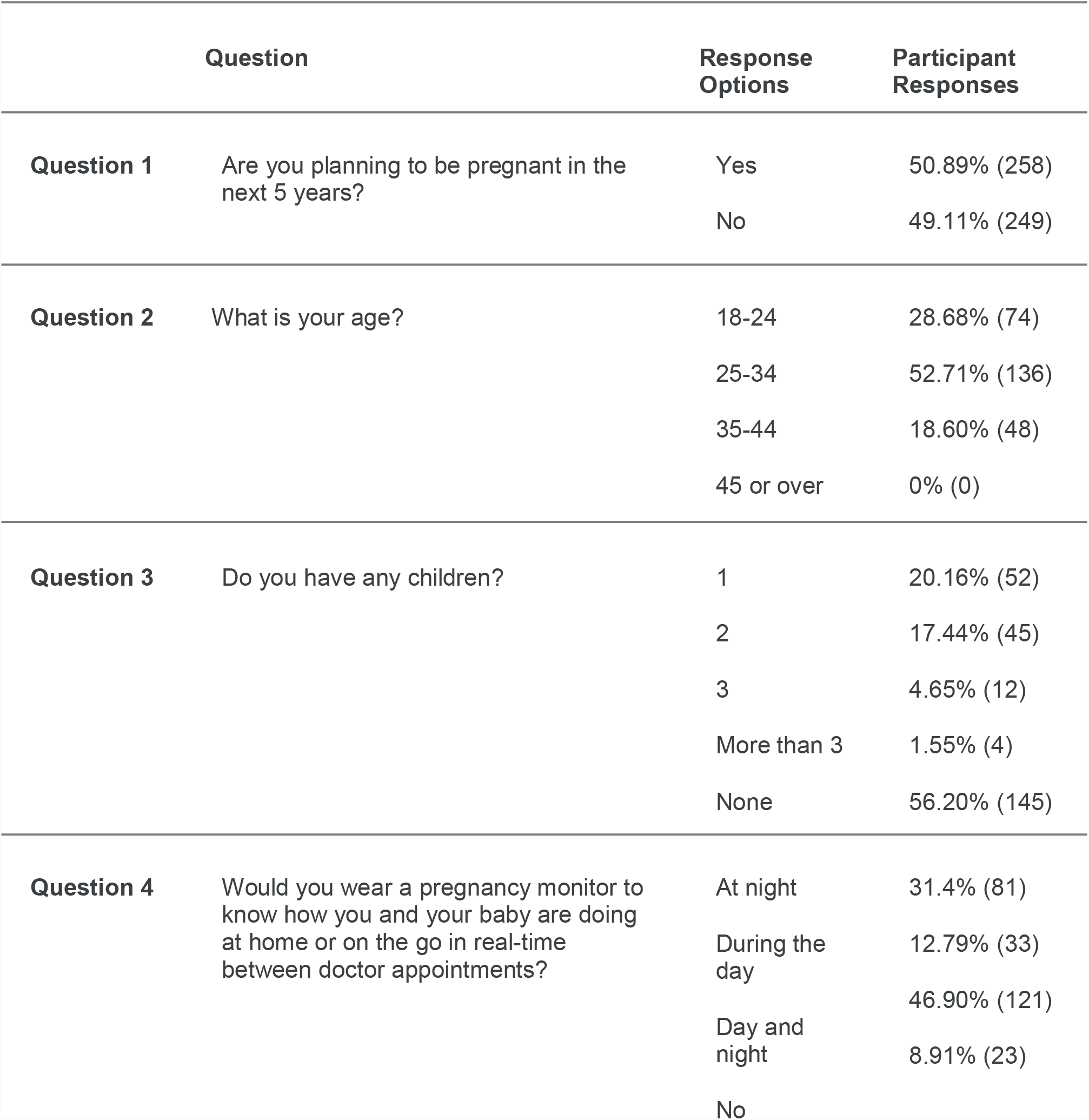
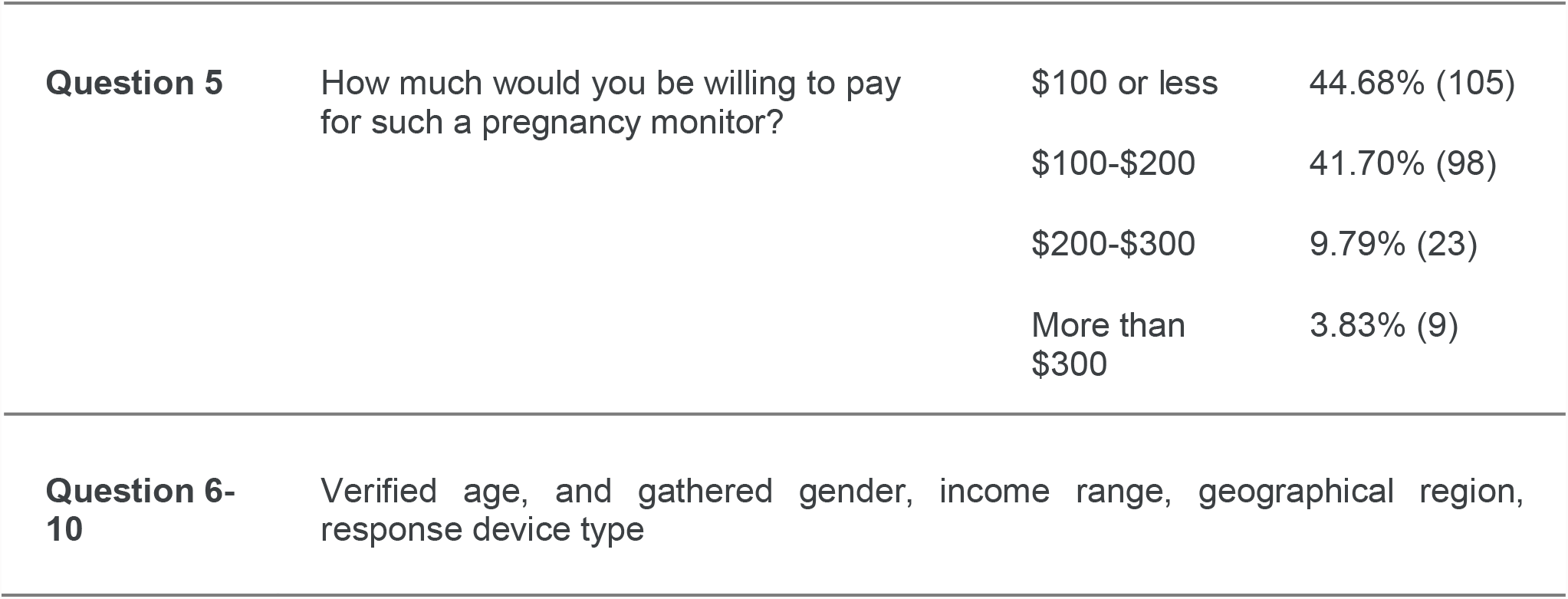
Survey results.

**Figure 3.**
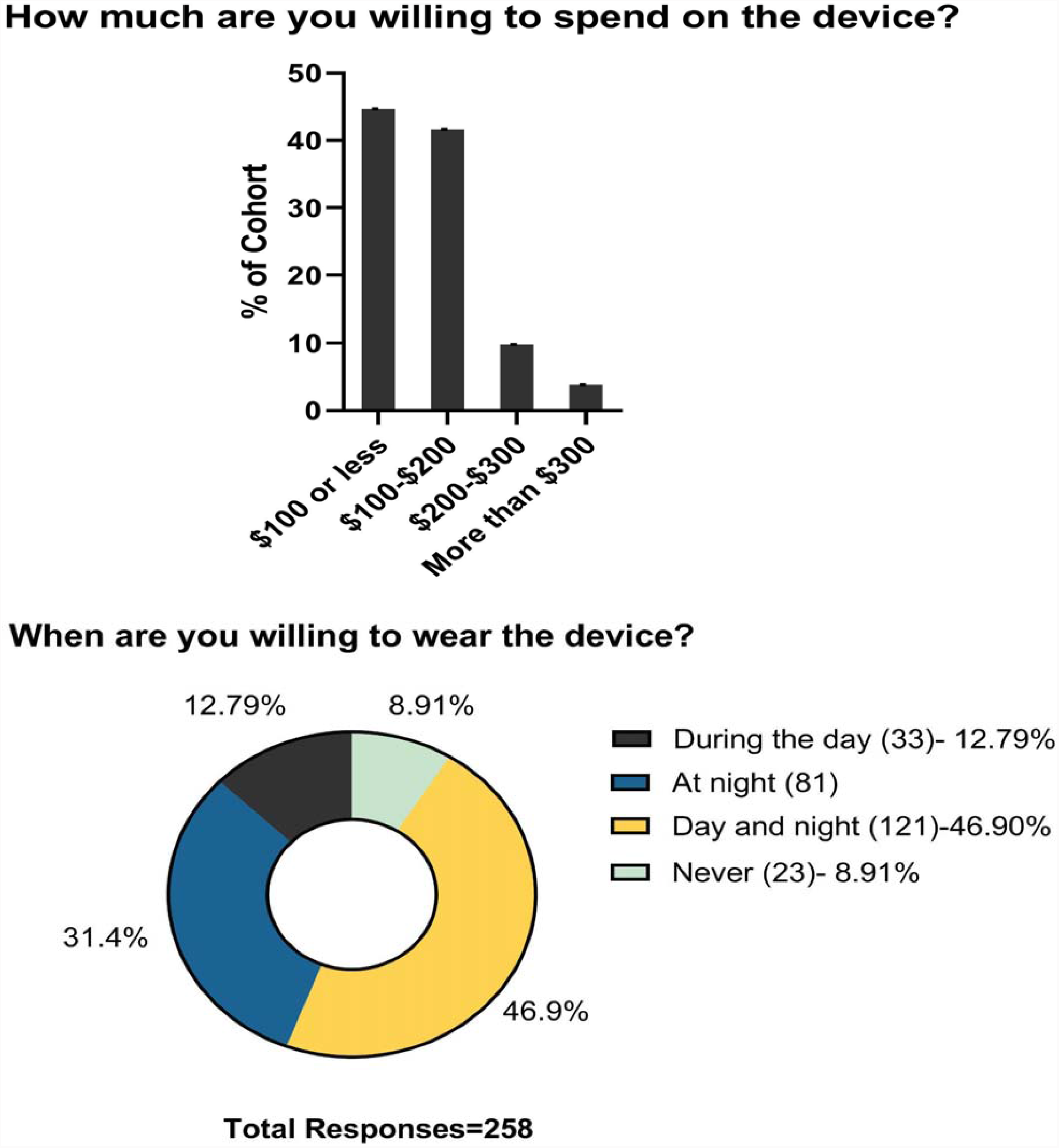
Survey findings.

## Discussion

### Main Findings

A cohort of 508 women of childbearing age, the largest population sample surveyed to date for this purpose, demonstrated an overwhelming enthusiasm towards wearable ECG devices for monitoring maternal-fetal health during pregnancy in-home, while asleep and on the go. These findings indicate that the public is ready for the integration of wearable continuous ECG monitoring into the modern healthcare model of telemedicine.

### Strengths and Limitations

The strengths of the present study include a large cross-sectional US-wide representative population sampling approach with a response rate over 90% and a margin of error around 4%. The key limitation is that we conducted no piloting and validation of the questions 4 and 5 deployed in our survey. A mitigating circumstance is the fact that the questions were either binary or simple multiple-choice queries leaving little room for interpretation.

### Interpretation

The time for disruption of the present antiquated maternal-fetal monitoring antepartum has arrived. The required hardware and software technologies have matured and converged with the people’s readiness for telemedicine throughout the antepartum period.

Wearable products that monitor adult health and fitness do exist but are not targeted specifically towards pregnant women. Therefore, currently, no technology exists that provides information on the pregnancy physiology or the health status of the fetus or the pregnant mother. Non-wearable products that monitor aspects of fetal health exist in the consumer space. However, such devices all share common pitfalls like low fidelity ultrasound technology, very short-term (seconds to minutes) observations without data recordings and provide minimal data analytics (if any) value beyond brief entertainment and possibly perilous false reassurance.

At the same time, the cost of hardware components required to build high-quality ECG devices for this purpose has come down to such a level that continuous antepartum monitoring technology is now achievable within the price range identified in the present study. All required software components are also available.^3,5–7^

From physiological viewpoint, a pregnant woman and her unborn baby are intimately connected forming one single physiological system. Consequently, important information about their joint wellbeing is lost when they are not monitored together.^8^ With one recent exception, all current devices use periodic ultrasound-based monitoring which is less precise than ECG-based monitoring and cannot provide the continuous monitoring (during maternal sleep for example) that is possible with ECG-based wearable devices.^9^

## Conclusion

ECG-based monitoring devices for pregnant women will enable deeper analyses and more precise reporting of health status than current ultrasound-based devices. This report shows that this is a technology that mothers want and suggests that the time is right for maternal and fetal medicine to integrate it into healthcare delivery.

## Ethics statement

Each survey participant was first given the option to opt out of the survey (Question 1). Only if agreed to continue, were the subsequent questions shown. Due to the completely anonymous nature of the data collection and the provided right to opt out of the survey or agree to continue, no IRB approval was obtained prior to the survey’s completion. However, after the completion of the survey and prior to publication of the findings, we consulted the UW IRB. It was determined that the approach taken meets the definition of “minor non-compliance” with IRB approved procedures/UW policies and procedures, because it posed no significant increase in risk or any decrease in benefits to subjects. The IRB ruled that this means that the data collected as part of this research cannot be described as part of a study reviewed by human subject division (IRB). On January 24, 2022, the IRB determined that the corrective actions described in the report are sufficient. No additional actions are required at this time.

## Supporting information

UW IRB ethics determination

## Data Availability

All data produced in the present study are available upon reasonable request to the authors.

## Acknowledgments

The authors gratefully acknowledge the assistance of D. Levitan and Y. Kimmelfeld during the survey design.

## References

1. Mann DM, Chen J, Chunara R, Testa PA, Nov O. COVID-19 transforms health care through telemedicine: Evidence from the field. J Am Med Inform Assoc [Internet]. 2020 Jul 1;27(7):1132–5. Available from: http://dx.doi.org/10.1093/jamia/ocaa072

2. Frasch MG. Letter to the Editor: Mind the gap: epistemology of heart rate variability. Am J Physiol Regul Integr Comp Physiol [Internet]. 2020 Sep 1;319(3):R343–4. Available from: http://dx.doi.org/10.1152/ajpregu.00183.2020

3. Sarkar P, Lobmaier S, Fabre B, González D, Mueller A, Frasch MG, et al. Detection of maternal and fetal stress from the electrocardiogram with self-supervised representation learning. Sci Rep [Internet]. 2021 Dec 17;11(1):24146. Available from: http://dx.doi.org/10.1038/s41598-021-03376-8

4. Lobmaier SM, Müller A, Zelgert C, Shen C, Su PC, Schmidt G, et al. Fetal heart rate variability responsiveness to maternal stress, non-invasively detected from maternal transabdominal ECG. Arch Gynecol Obstet [Internet]. 2020 Feb;301(2):405–14. Available from: http://dx.doi.org/10.1007/s00404-019-05390-8

5. Su L, Wu H-T. Extract Fetal ECG from Single-Lead Abdominal ECG by De-Shape Short Time Fourier Transform and Nonlocal Median. sFrontiers in Applied Mathematics and Statistics [Internet]. 2017;3:2. Available from: https://www.frontiersin.org/article/10.3389/fams.2017.00002

6. Frasch MG. Heart rate variability code: Does it exist and can we hack it? [Internet]. arXiv [q-bio.TO]. 2020. Available from: http://arxiv.org/abs/2001.08264

7. Frasch MG. Comprehensive HRV estimation pipeline using Neurokit2 [Internet]. Available from: https://zenodo.org/badge/latestdoi/427111001

8. Frasch MG, Lobmaier SM, Stampalija T, Desplats P, Pallarés ME, Pastor V, et al. Non-invasive biomarkers of fetal brain development reflecting prenatal stress: An integrative multi-scale multi-species perspective on data collection and analysis. Neurosci Biobehav Rev [Internet]. 2018 May 30; Available from: http://dx.doi.org/10.1016/j.neubiorev.2018.05.026

9. Mhajna M, Schwartz N, Levit-Rosen L, Warsof S, Lipschuetz M, Jakobs M, et al. Wireless, remote solution for home fetal and maternal heart rate monitoring. American Journal of Obstetrics & Gynecology MFM [Internet]. 2020 Mar 17;100101. Available from: http://www.sciencedirect.com/science/article/pii/S2589933320300318

