## Supplementary material for "Wearable technology for health monitoring during pregnancy: an observational cross-sectional survey study": UW IRB ethics determination

### REVIEW OF REPORTABLE NEW INFORMATION

January 24, 2022

Dear Martin G Frasch:

This letter is in regard to:

| | |
|---|---|
| Type of Review: | Reportable New Information |
| Title: | ECG wearables during pregnancy |
| Submitted by: | Martin G Frasch |
| Responsible Party: | Martin G Frasch |
| IRB ID: | RNI00001741 |

**RNI DETERMINATION**

On January 21, 2022, the Human Subjects Division reviewed the following Reportable New Information:

The RNI explained that you conducted an online cross-sectional survey-based study using a SurveyMonkey audience with 507 individuals between July 31, 2019 to August 1, 2019. You explained that SurveyMonkey is an online survey tool used to collect data from individuals across the US population. You further explained that a pool of over two million people is maintained through an agreement wherein participants agree to take part in a survey, in exchange for SurveyMonkey donating $0.50 to the charity of the individual's choice. The SurveyMonkey algorithm then randomly assigns participants to surveys in a manner that creates a sample representation of the demographics specified by the researcher.

It was your understanding that based on the information from the SurveyMonkey Audience, that no IRB process was required. However, you now wish to publish the data, but it will not be accepted without IRB approval or acknowledgement from the IRB.

The reported problem meets the definition of non-compliance with IRB approved procedures/UW policies and procedures, but it is minor non-compliance because it posed no significant increase in risk or any decrease in benefits to subjects.

Please note that as Human subjects research, as defined by federal regulations under 45 CFR 46.102, was conducted prior to HSD review and determination of exempt status. Please be advised that our office cannot provide retroactive approval for research conducted without IRB approval. This means that the data collected as part of your research cannot be described (for example, in publication) as being part of a study reviewed by HSD.

**IRB REVIEW**

On January 24, 2022, the IRB determined that:

- The corrective actions described in the report are sufficient. No additional actions are required at this time.

Please do not hesitate to contact me if you have any questions.

Sincerely,

Gloria Park, Compliance Administrator
